## Supplementary Information for "Associations of proton pump inhibitors with susceptibility to influenza, pneumonia, and COVID-19: Evidence from a large population based cohort study"

**Supplementary Figure 1.** Directed acyclic graph (DAG) for evaluation of covariates in the logistic regression model.

**Supplementary Figure 2.** Kaplan-Meier curves illustrating the event-free probability for the outcomes among users and non-users of proton pump inhibitors (PPIs).

**Supplementary Figure 3.** Stratified analysis of proton pump inhibitor (PPI) users and the risk of COVID-19 severity and mortality.

**Supplementary Figure 4.** Directed acyclic graph (DAG) for potential unmeasured confounders.

**Supplementary Table S1.** Generic name and examples of trade name of proton pump inhibitors.

**Supplementary Table S2.** Definitions of outcomes in the UK Biobank cohort.

**Supplementary Table S3.** The proportional hazards assumption tested by Schoenfeld residuals tests.

**Supplementary Table S4.** Associations of PPI use with the risk of influenza, pneumonia, and other respiratory infections (with inclusion of self-reported cases).

**Supplementary Table S5.** Associations of PPI use with COVID-19 severity and mortality.

**Supplementary Table S6.** Associations of PPI use with the risk of influenza, pneumonia, COVID-19, and other respiratory infections by different types of PPIs.

**Supplementary Table S7.** Associations of PPI use with the risk of influenza, pneumonia, COVID-19, and other respiratory infections by *CYP2C19* phenotypes

**Supplementary Table S8.** Associations of PPI use with COVID-19 severity and mortality by *CYP2C19* phenotypes

**Supplementary Table S9.** Associations of PPI use with the risk of influenza, pneumonia, COVID-19, and other respiratory infections with multiple imputation.

**Supplementary Table S10.** Analysis of associations of PPI use with COVID-19 severity and mortality with multiple imputation.

**Supplementary Table S11.** Clinical characteristics of included participants after propensity score-matching.

**Supplementary Table S12.** Propensity score-matched analysis of associations of PPI use with the risk of influenza, pneumonia, COVID-19, and other respiratory infections.

**Supplementary Table S13.** Propensity score-matched analysis of associations of PPI use with COVID-19 severity and mortality.

**Supplementary Table S14.** Comparisons between proton pump inhibitor (PPI) and histamine-2 receptor antagonist (H2RA) users for COVID-19 severity and mortality.

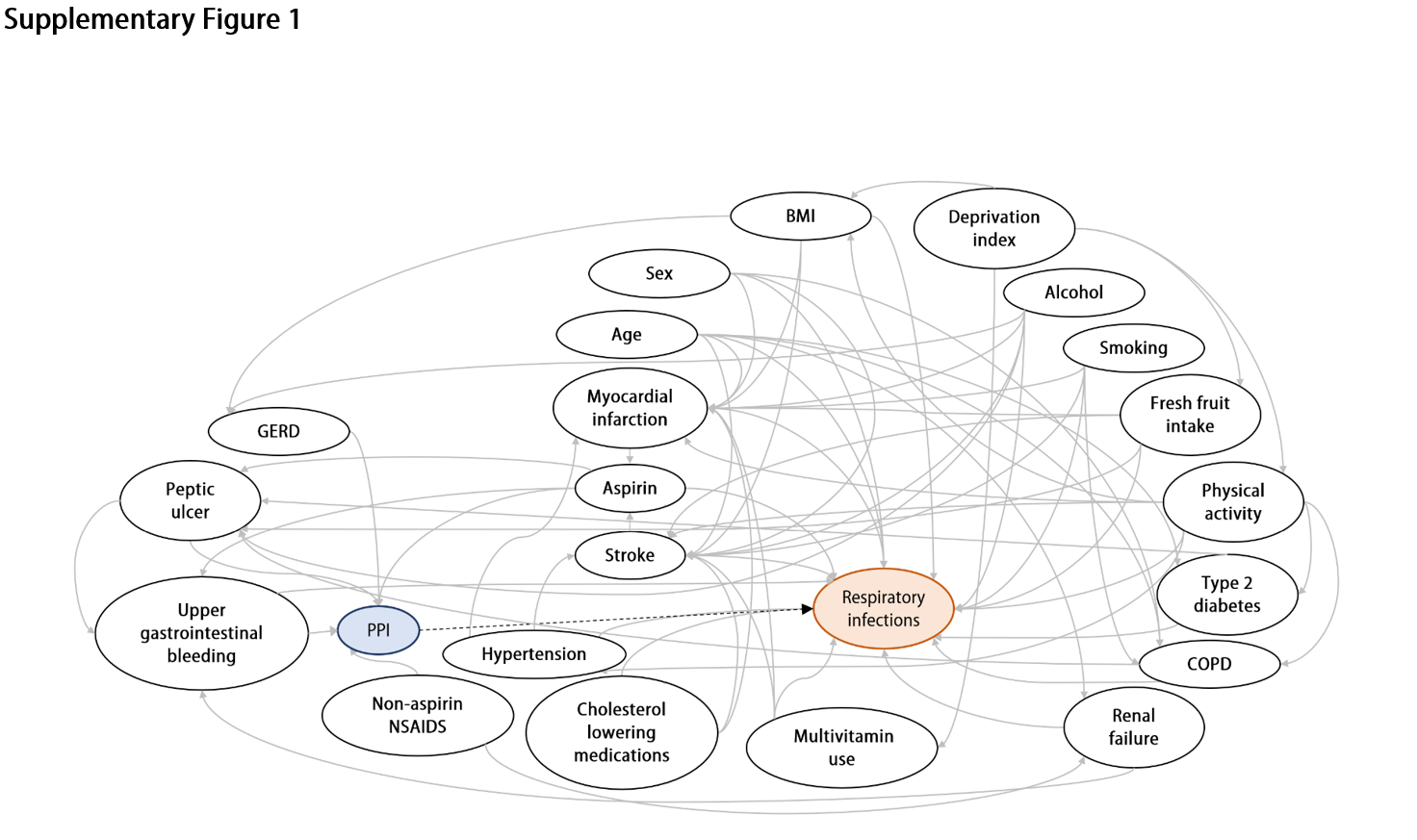

**Supplementary Figure 1.** **Directed acyclic graph (DAG) for evaluation of covariates in the logistic regression model.** Nodes and arrows represent variables and causal associations, respectively. The exposure (proton pump inhibitor, PPI) is denoted by a blue node, and the outcome (respiratory infections) is labeled by an orange node. The proposed causal associations were based on existed literature or expert knowledge. BMI: body mass index; COPD: chronic obstructive pulmonary disease; GERD: gastroesophageal reflux disease; H2RA: histamine-2 receptor antagonist; PPI: proton pump inhibitor.

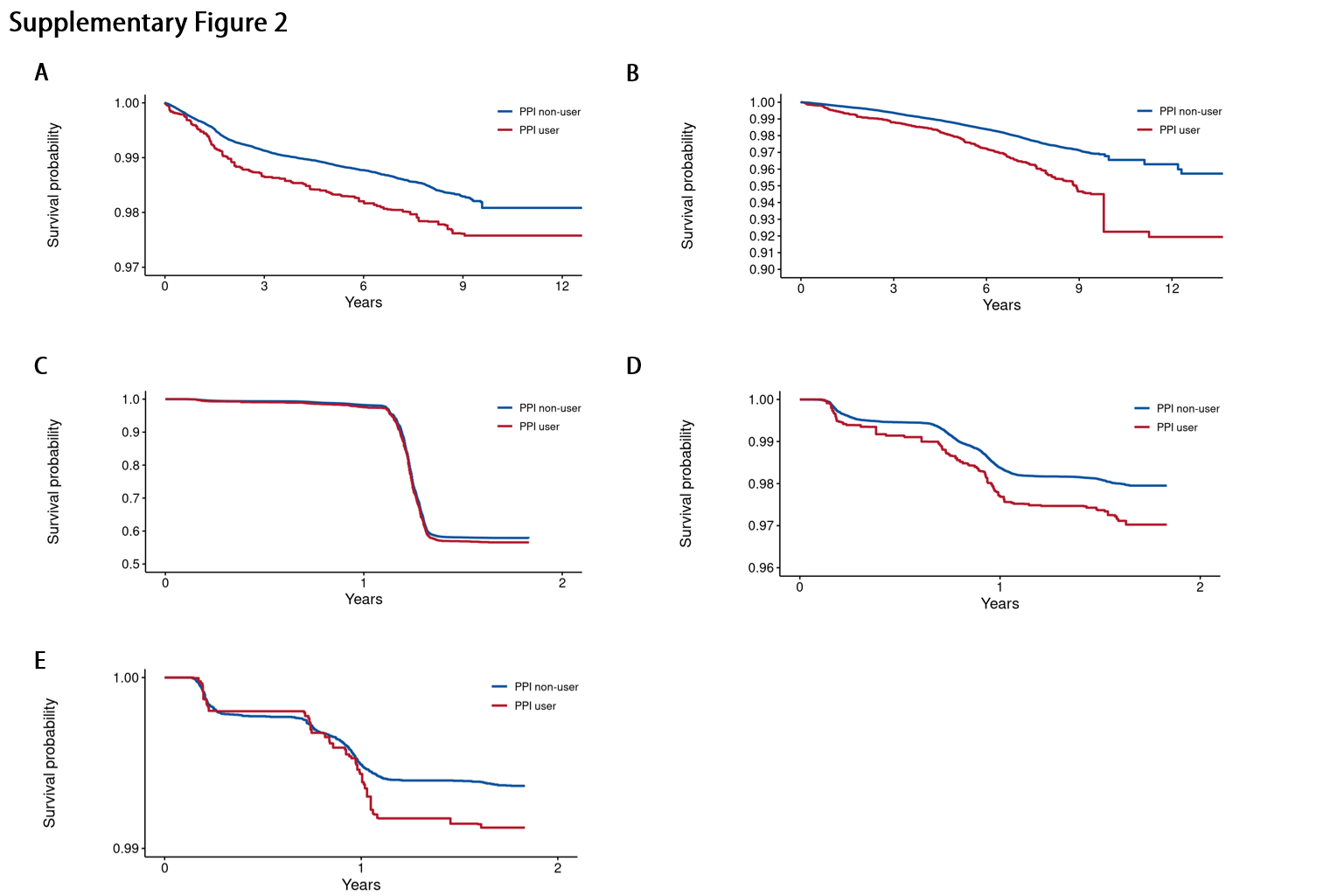

**Supplementary Figure 2. Inverse probability weights-adjusted** **Kaplan-Meier curves illustrating the event-free probability for the outcomes among users and non-users of proton pump inhibitors (PPIs).** **A.** Influenza. **B.** Pneumonia. **C.** COVID-19 positivity. **D.** COVID-19 severity. **E.** COVID-19 mortality.

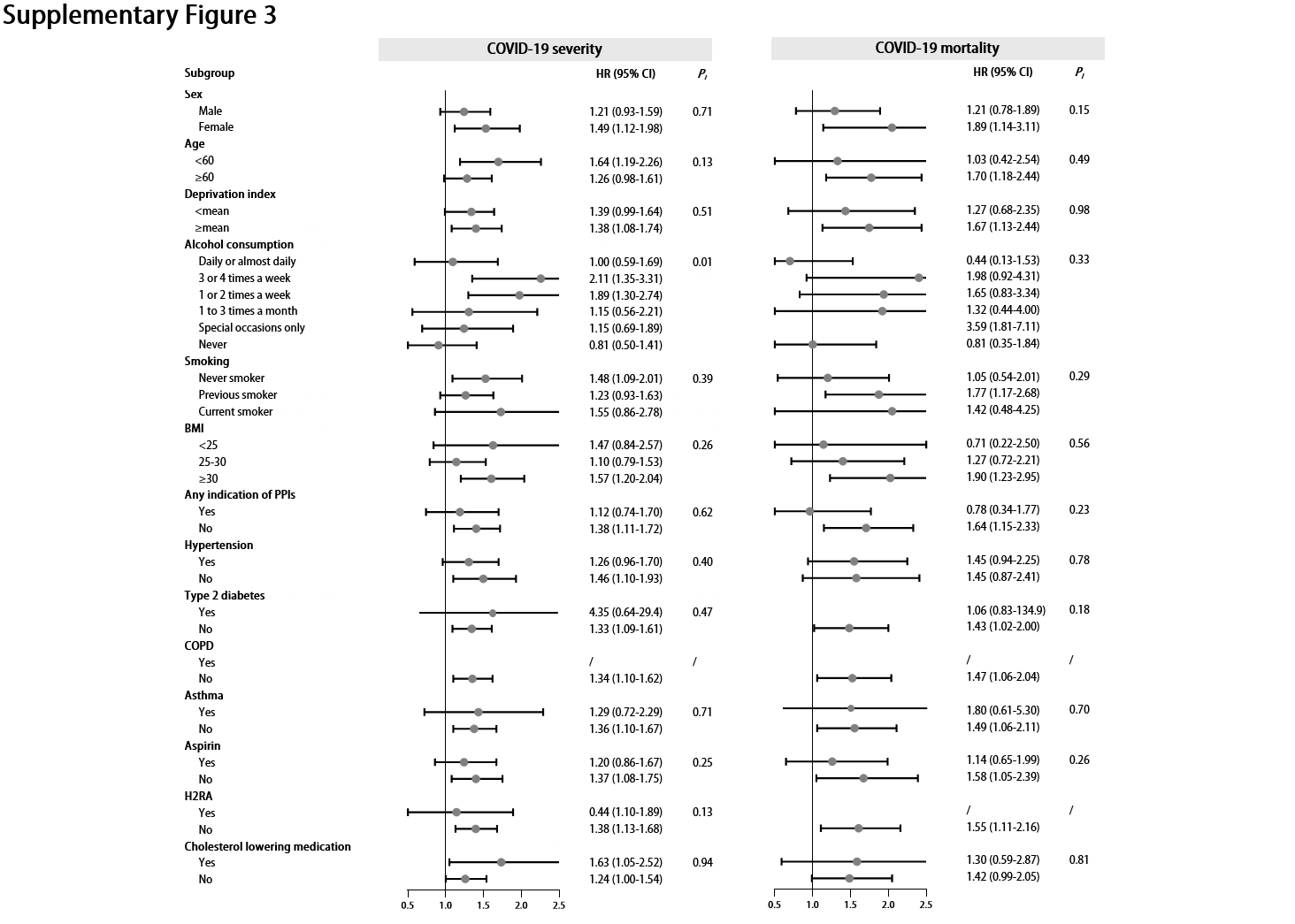

**Supplementary Figure 3. Stratified analysis of proton pump inhibitor (PPI) users and the risk of COVID-19 severity and mortality.** Effect estimates were based on age, sex, deprivation index, alcohol consumption, smoking, body mass index (BMI), indications of PPIs, hypertension, type 2 diabetes, chronic obstructive pulmonary disease (COPD), asthma, aspirin, histamine 2 receptor antagonist (H2RA), and cholesterol-lowering medication, using the fully adjusted model. CI: confidence interval; HR: hazard ratio; P_i_: *P* value for interaction.

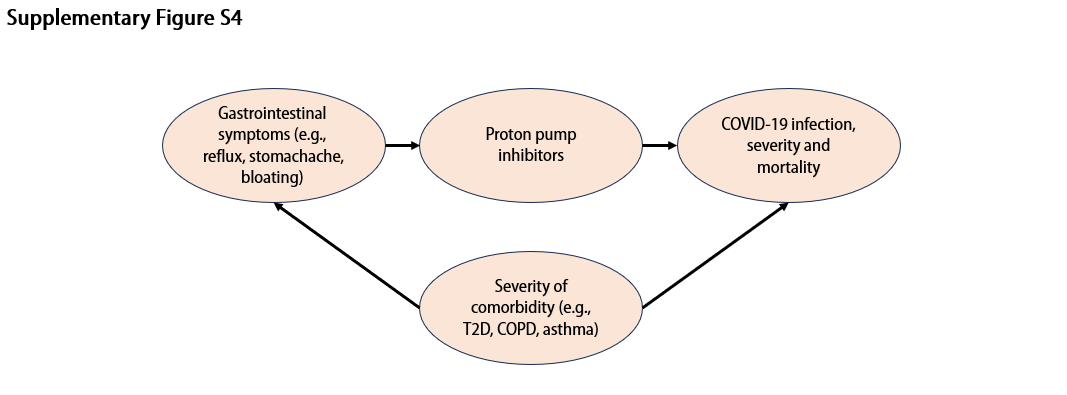

**Supplementary Figure 4.** Directed acyclic graph (DAG) for potential unmeasured confounders. Nodes and arrows represent variables and causal associations, respectively. COPD: chronic obstructive pulmonary disease; COVID: coronavirus disease; T2D: type 2 diabetes.

**Supplementary Table S1.** Generic name and examples of trade name of proton pump inhibitors.

| **Generic name** | **Trade name** |
| --- | --- |
| [Dexlansoprazole](https://www.drugs.com/mtm/dexlansoprazole.html) | [Dexilant](https://www.drugs.com/mtm/dexilant.html) |
| [Esomeprazole](https://www.drugs.com/mtm/esomeprazole.html) | [Nexium](https://www.drugs.com/nexium.html) |
| [Lansoprazole](https://www.drugs.com/lansoprazole.html) | [Prevacid](https://www.drugs.com/prevacid.html) |
| [Omeprazole](https://www.drugs.com/omeprazole.html) | [Prilosec](https://www.drugs.com/prilosec.html) |
| [Pantoprazole](https://www.drugs.com/pantoprazole.html) | [Protonix](https://www.drugs.com/protonix.html) |
| [Rabeprazole](https://www.drugs.com/mtm/rabeprazole.html) | [Aciphex](https://www.drugs.com/aciphex.html) |

Data retrieved from http://www.drugs.com.

**Supplementary Table S1.** Definitions of outcomes in the UK Biobank cohort.

| **Outcome** | **Description of outcome*** |
| --- | --- |
| **Influenza** | Influenza due to certain identified influenza virus |
|  | Influenza due to identified influenza virus |
|  | Influenza, virus not identified |
| **Pneumonia** | Viral pneumonia, not elsewhere classified |
|  | Pneumonia due to streptococcus pneumoniae |
|  | Pneumonia due to haemophilus influenzae |
|  | Bacterial pneumonia, not elsewhere classified |
|  | Pneumonia due to other infectious organisms, not elsewhere classified |
|  | Pneumonia in diseases classified elsewhere |
|  | Pneumonia, organism unspecified |
| **COVID-19 infection** | Positive COVID test, and included patients with in-patient COVID diagnosis and cause of death due to COVID-19 |
| **COVID-19 severity** | In-patient diagnosis of COVID-19 |
| **COVID-19 mortality** | Underlying (primary) cause of death due to COVID-19 |
| **Other upper respiratory infections** | Acute nasopharyngitis (common cold) |
|  | Acute sinusitis |
|  | Acute pharyngitis |
|  | Acute tonsillitis |
|  | Acute laryngitis and tracheitis |
|  | Acute obstructive laryngitis (croup) and epiglottitis |
|  | Acute upper respiratory infections of multiple and unspecified sites |
| **Other lower respiratory infections** | Acute bronchitis |
|  | Acute bronchiolitis |
|  | Unspecified acute lower respiratory infection |

*First reported date was used for the outcome.

**Supplementary Table S2.** The proportional hazards assumption tested by Schoenfeld residuals tests.

| **Outcome** | **Schoenfeld residuals test (*P*-value)** |
| --- | --- |
| **Influenza** | 0.428 |
| **Pneumonia** | 0.766 |
| **COVID-19 infection** | 0.054 |
| **COVID-19 severity** | 0.347 |
| **COVID-19 mortality** | 0.430 |
| **Other upper respiratory infections** | 0.253 |
| **Other lower respiratory infections** | 0.372 |

**Supplementary Table S3.** Associations of PPI use with the risk of influenza, pneumonia, and other respiratory infections (with inclusion of self-reported cases).

|  | **Cases / Person-years** | **HR (95% CI)*** | ***P*** |
| --- | --- | --- | --- |
| **Influenza** |  |  |  |
| **Non-regular PPI use** | 2 013/6 051 | 1.00 (Reference) | **0.001** |
| **Regular PPI use** | 184/546 | 1.32 (1.12-1.56) |  |
| **Pneumonia** |  |  |  |
| **Non-regular PPI use** | 2 917/12 916 | 1.00 (Reference) |  |
| **Regular PPI use** | 380/1 711 | 1.42 (1.26-1.60) | **<0.001** |
| **Other upper respiratory infection** |  |  |  |
| **Non-regular PPI use** | 14 403/52 344 | 1.00 (Reference) |  |
| **Regular PPI use** | 1 119/3 990 | 1.19(1.17-1.28) | **<0.001** |
| **Other lower respiratory infection** |  |  |  |
| **Non-regular PPI use** | 14 498/55 414 | 1.00 (Reference) |  |
| **Regular PPI use** | 1 487/5 604 | 1.38 (1.30-1.46) | **<0.001** |

*Adjusted for age, sex, ethnicity, deprivation index, smoking, alcohol consumption, physical activity, fresh fruit intake, body mass index, any indication of PPIs (gastroesophageal reflux disease [GERD], peptic ulcer, upper gastrointestinal bleeding), comorbidities (hypertension, type 2 diabetes, renal failure, myocardial infarction, stroke, chronic obstructive pulmonary disease [COPD], asthma), medications (aspirin, non-aspirin non-steroidal anti-inflammatory drugs [NSAIDs, ibuprofen], histamine 2 receptor antagonists (H2RAs), cholesterol lowering medications), multivitamin use, and influenza vaccination (for influenza).

CI: confidence interval; COVID-19: coronavirus disease 2019; HR: hazard ratio; PPI: proton pump inhibitor.

**Supplementary Table S4.** Associations of PPI use with COVID-19 severity and mortality.

|  | **Case/person-years** | **Non-adjusted model** | | **Age/sex-adjusted model** | | **Fully adjusted model*** | |
| --- | --- | --- | --- | --- | --- | --- | --- |
|  |  | **HR (95% CI)** | ***P*** | **HR (95% CI)** | ***P*** | **HR (95% CI)** | ***P*** |
| **COVID-19 severity** |  |  |  |  |  |  |  |
| **Non-regular PPI use** | 1 104/844 | 1.00 (reference) |  | 1.00 (reference) |  | 1.00 (reference) |  |
| **Regular PPI use** | 138/100 | 2.32 (1.94-2.77) | **<0.001** | 1.95 (1.64-2.34) | **<0.001** | 1.33 (1.09-1.61) | **0.004** |
| **COVID-19 mortality** |  |  |  |  |  |  |  |
| **Non-regular PPI use** | 337/239 | 1.00 (reference) |  | 1.00 (reference) |  | 1.00 (reference) |  |
| **Regular PPI use** | 48/36 | 2.61 (1.93-3.54) | **<0.001** | 1.90 (1.40-2.58) | **<0.001** | 1.46(1.05-2.03) | **0.024** |

*Adjusted for age, sex, ethnicity, deprivation index, smoking, alcohol consumption, physical activity, fresh fruit intake, body mass index, any indication of PPIs (gastroesophageal reflux disease [GERD], peptic ulcer, upper gastrointestinal bleeding), comorbidities (hypertension, type 2 diabetes, renal failure, myocardial infarction, stroke, chronic obstructive pulmonary disease [COPD], asthma), medications (aspirin, non-aspirin non-steroidal anti-inflammatory drugs [NSAIDs, ibuprofen], histamine 2 receptor antagonists (H2RAs), cholesterol lowering medications), multivitamin use, and COVID-19 vaccination .

CI: confidence interval; COVID-19: coronavirus disease 2019; HR: hazard ratio; PPI: proton pump inhibitor.

**Supplementary Table S5.** Associations of PPI use with the risk of influenza, pneumonia, COVID-19, and other respiratory infections by different types of PPIs.

|  | **Case/Person-years** | **HR (95% Cl)*** | ***P*** |
| --- | --- | --- | --- |
| **Influenza** |  |  |  |
| Non-regular PPI use | 2 009/6 011 | 1.00 (Reference) |  |
| Omeprazole | 96/306 | 1,36 (1.09-1.70) | **0.006** |
| Lansoprazole | 33/75 | 0.92 (0.77-1.10） | 0.371 |
| Esomeprazole | 1/1 | 0.64 (0.33-1.24) | 0.186 |
| Rabeprazole | 1/1 | 0.87 (0.53-1,42) | 0.570 |
| Pantoprazole | **-** | **-** | **-** |
| **Pneumonia** |  |  |  |
| Non-regular PPI use | 2 904/12 867 | 1.00 (Reference) |  |
| Omeprazole | 4 193/30 701 | 1.49 (1.29-1.74) | **<0.001** |
| Lansoprazole | 2 459/17 382 | 1.54 (1.27-1.86) | **<0.001** |
| Esomeprazole | 233/1 730 | 1.66 (0.96-2.88) | 0.069 |
| Rabeprazole | 118/868 | 1.10 (0.46-2.66) | 0.827 |
| Pantoprazole | 143/1 070 | 1.58 (0.78-3.17) | 0.201 |
| **Other upper respiratory infection** |  |  |  |
| Non-regular PPI use | 14 449/52 499 | 1.00 (Reference) |  |
| Omeprazole | 534/1 959 | 1.14 (1.04-1.25) | **0.006** |
| Lansoprazole | 411/1 085 | 1.18 (1.05-1.33) | **0.006** |
| Esomeprazole | 28/95 | 1.17 (0.81-1.70) | 0.408 |
| Rabeprazole | 9/38 | 0.65 (0.34-1.25) | 0.193 |
| Pantoprazole | 21/74 | 1.41 (0.92-2.16) | 0.119 |
| **Other lower respiratory infection** |  |  |  |
| Non-regular PPI use | 14 494/55 384 | 1.00 (Reference) |  |
| Omeprazole | 705/2 723 | 1.31 (1.21,1.42) | **<0.001** |
| Lansoprazole | 397/1 424 | 1.30 (1.17,1.44) | **<0.001** |
| Esomeprazole | 39/148 | 1.35 (0.98,1.85) | 0.066 |
| Rabeprazole | 16/67 | 0.94 (0.58,1.54) | 0.813 |
| Pantoprazole | 23/107 | 1.23 (0.82,1.86) | 0.317 |
| **COVID-19 positivity** |  |  |  |
| Non-regular PPI use | 23 989/29 080 | 1.00 (Reference) |  |
| Omeprazole | 1 347/1 585 | 0.99 (0.88,1.12) | 0.927 |
| Lansoprazole | 769/912 | 0.89 (0.77,1.04) | 0.151 |
| Esomeprazole | 65/78 | 1.07 (0.63,1.78) | 0.801 |
| Rabeprazole | 33/41 | 1.16 (0.60,2.27) | 0.663 |
| Pantoprazole | 44/53 | 0.99 (0.88,1.12) | 0.853 |
| **COVID-19 severity** |  |  |  |
| Non-regular PPI use | 1 104/844 | 1.00 (Reference) |  |
| Omeprazole | 137/95 | 1.34 (1.10,1.63) | **0.003** |
| Lansoprazole | 70 /51 | 1.20 (0.92,1.55) | 0.173 |
| Esomeprazole | 4/3 | 0.69 (0.26,1.85) | 0.459 |
| Rabeprazole | 5/6 | 1.68 (0.69,4.09） | 0.249 |
| Pantoprazole | 1/1 | 0.35 (0.05,2.48) | 0.291 |
| **COVID-19 mortality** |  |  |  |
| Non-regular PPI use | 337/239 | 1.00 (Reference) |  |
| Omeprazole | 49/36 | 1.48 (1.07,2.05) | **0.018** |
| Lansoprazole | 19/15 | 0.99 (0.77,1.27) | 0.933 |
| Esomeprazole | 3/3 | 1.83 (0.58,5.80) | 0.303 |
| Rabeprazole | - | - | - |
| Pantoprazole | 3/2 | 3.77 (1.18,12.06) | 0.025 |

*Adjusted for age, sex, ethnicity, deprivation index, smoking, alcohol consumption, physical activity, fresh fruit intake, body mass index, any indication of PPIs (gastroesophageal reflux disease [GERD], peptic ulcer, upper gastrointestinal bleeding), comorbidities (hypertension, type 2 diabetes, renal failure, myocardial infarction, stroke, chronic obstructive pulmonary disease [COPD], asthma), medications (aspirin, non-aspirin non-steroidal anti-inflammatory drugs [NSAIDs, ibuprofen], histamine 2 receptor antagonists (H2RAs), cholesterol lowering medications), multivitamin use, and influenza vaccination (for influenza) or COVID-19 vaccination (for COVID-19-related outcomes).

CI: confidence interval; COVID-19: coronavirus disease 2019; HR: hazard ratio; PPI: proton pump inhibitor.

**Supplementary Table S6.** Associations of PPI use with the risk of influenza, pneumonia, COVID-19, and other respiratory infections by *CYP2C19* phenotypes

|  | **Case/Person-years** | **HR (95% Cl)*** | ***P*** |
| --- | --- | --- | --- |
| **Influenza** |  |  |  |
| Non-regular PPI user | 2 009/6 011 | 1.00 (reference) |  |
| PPI user, *CYP2C19* rapid and ultrarapid metabolizers | 68/193 | 1.27 (0.98-1.64) | 0.066 |
| PPI user, *CYP2C19* normal metabolizers | 94/285 | 1.34 (1.07-1.67) | **0.010** |
| PPI user, *CYP2C19* likely intermediate, intermediate and poor metabolizers | 5/17 | 1.67 (0.69-4.03) | 0.257 |
| **Pneumonia** |  |  |  |
| Non-regular PPI user | 2 904/12 867 | 1.00 (reference) |  |
| PPI user, *CYP2C19* rapid and ultrarapid metabolizers | 146/626 | 1.45 (1.22-1.73) | **<0.001** |
| PPI user, *CYP2C19* normal metabolizers | 186/869 | 1.32 (1.13-1.55) | **<0.001** |
| PPI user, *CYP2C19* likely intermediate, intermediate and poor metabolizers | 2/14 | 1.24 (0.98-1.56) | 0.078 |
| **COVID-19** |  |  |  |
| Non-regular PPI user | 23 989/29 080 | 1.00 (reference) |  |
| PPI user, *CYP2C19* rapid and ultrarapid metabolizers | 516/607 | 1.11 (0.97,1.27) | 0.144 |
| PPI user, *CYP2C19* normal metabolizers | 769/910 | 1.08 (0.96.1.21) | 0.184 |
| PPI user, *CYP2C19* likely intermediate, intermediate and poor metabolizers | 32/36 | 1.22 (0.71,2.12) | 0.469 |
| **Other upper respiratory infection** |  |  |  |
| Non-regular PPI user | 14 449/52 499 | 1.00 (reference) |  |
| PPI user, *CYP2C19* rapid and ultrarapid metabolizers | 392/1 416 | 1.10 (0.99-1.22) | 0.078 |
| PPI user, *CYP2C19* normal metabolizers | 609/2 151 | 1.26 (1.15-1.37) | **<0.001** |
| PPI user, *CYP2C19* likely intermediate, intermediate and poor metabolizers | 14/54 | 0.70 (0.42-1.19) | 0.190 |
| **Other lower respiratory infection** |  |  |  |
| Non-regular PPI user | 14 494/55 384 | 1.00 (reference) |  |
| PPI user, *CYP2C19* rapid and ultrarapid metabolizers | 546/2 049 | 1.32 (1.21-1.45) | **<0.001** |
| PPI user, *CYP2C19* normal metabolizers | 763/2 868 | 1.36 (1.26-1.47) | **<0.001** |
| PPI user, *CYP2C19* likely intermediate, intermediate and poor metabolizers | 26/104 | 1.19 (0.81-1.75) | 0.379 |

CI: confidence interval; COVID-19: coronavirus disease 2019; HR: hazard ratio; Pi: *P* for interaction among different types of metabolizers; PPI: proton pump inhibitor.

*Fully adjusted model

**Supplementary Table S7.** Associations of PPI use with COVID-19 severity and mortality by *CYP2C19* phenotypes

|  | **Case/Person-years** | **HR (95% Cl)** | ***P*** |
| --- | --- | --- | --- |
| **COVID-19 severity** |  |  |  |
| Non-regular PPI user | 1 104/844 | 1.00 (reference) |  |
| PPI user, *CYP2C19* Rapid and ultrarapid metabolizers | 48/32 | 1.33 (0.98,1.80) | 0.065 |
| PPI user, *CYP2C19* normal metabolizers | 77/475 | 1.01 (0.91,1.11) | 0.862 |
| PPI user, *CYP2C19* likely intermediate, intermediate and poor metabolizers | 8/7 | 4.45 (2.19,9.05) | **<0.001** |
| **COVID-19 mortality** |  |  |  |
| Non-regular PPI user | 337/239 | 1.00 (reference) |  |
| PPI user, *CYP2C19* Rapid and ultrarapid metabolizers | 20/15 | 1.81 (1.12,2.91) | **0.015** |
| PPI user, *CYP2C19* normal metabolizers | 23/16 | 1.26 (0.81,1.97) | 0.310 |
| PPI user, *CYP2C19* likely intermediate, intermediate and poor metabolizers | 3/3 | 5.78 (1.79,18.66) | **0.003** |

CI: confidence interval; COVID-19: coronavirus disease 2019; HR: hazard ratio; *P* for interaction among different types of metabolizers; PPI: proton pump inhibitor.

**Supplementary Table S8.** Associations of PPI use with the risk of influenza, pneumonia, COVID-19, and other respiratory infections with multiple imputation.

|  | **Cases / Person-years** | **HR (95% CI)*** | ***P*** |
| --- | --- | --- | --- |
| **Influenza** |  |  |  |
| **Non-regular PPI use** | 2 785/8 228 | 1.00 (Reference) |  |
| **Regular PPI use** | 251/745 | 1.29 (1.09-1.53) | **0.003** |
| **Pneumonia** |  |  |  |
| **Non-regular PPI use** | 3 964/17 479 | 1.00 (Reference) |  |
| **Regular PPI use** | 568/2 600 | 1.41 (1.28-1.55) | **<0.001** |
| **COVID-19 infection** |  |  |  |
| **Non-regular PPI use** | 80 134/96 108 | 1.00 (Reference) |  |
| **Regular PPI use** | 4 708/5 541 | 1.10 (1.05-1.15) | **<0.001** |
| **Other upper respiratory infection** |  |  |  |
| **Non-regular PPI use** | 20 125/72 592 | 1.00 (Reference) |  |
| **Regular PPI use** | 1 625/5 881 | 1.21 (1.14-1.28) | **<0.001** |
| **Other lower respiratory infection** |  |  |  |
| **Non-regular PPI use** | 20 403/77 850 | 1.00 (Reference) |  |
| **Regular PPI use** | 2 129/8 053 | 1.35 (1.28-1.42) | **<0.001** |

*Adjusted for age, sex, ethnicity, deprivation index, smoking, alcohol consumption, physical activity, fresh fruit intake, body mass index, any indication of PPIs (gastroesophageal reflux disease [GERD], peptic ulcer, upper gastrointestinal bleeding), comorbidities (hypertension, type 2 diabetes, renal failure, myocardial infarction, stroke, chronic obstructive pulmonary disease [COPD], asthma), medications (aspirin, non-aspirin non-steroidal anti-inflammatory drugs [NSAIDs, ibuprofen], histamine 2 receptor antagonists (H2RAs), cholesterol lowering medications), multivitamin use, and influenza vaccination (for influenza) or COVID-19 vaccination (for COVID-19-related outcomes).

CI: confidence interval; COVID-19: coronavirus disease 2019; HR: hazard ratio; PPI: proton pump inhibitor.

**Supplementary Table S9.** Analysis of associations of PPI use with COVID-19 severity and mortality with multiple imputation.

|  | **Cases / Person-years** | **HR (95% CI)*** | ***P*** |
| --- | --- | --- | --- |
| **COVID-19 severity** |  |  |  |
| **Non-regular PPI use** | 1 569/2 368 | 1.00 (Reference) |  |
| **Regular PPI use** | 202/18 309 | 1.47 (1.33,1.64) | **<0.001** |
| **COVID-19 mortality** |  |  |  |
| **Non-regular PPI use** | 2 858/43 802 | 1.00 (Reference) |  |
| **Regular PPI use** | 470/4 184 | 1.53 (1.27,1.86) | **<0.001** |

*Adjusted for age, sex, ethnicity, deprivation index, smoking, alcohol consumption, physical activity, fresh fruit intake, body mass index, any indication of PPIs (gastroesophageal reflux disease [GERD], peptic ulcer, upper gastrointestinal bleeding), comorbidities (hypertension, type 2 diabetes, renal failure, myocardial infarction, stroke, chronic obstructive pulmonary disease [COPD], asthma), medications (aspirin, non-aspirin non-steroidal anti-inflammatory drugs [NSAIDs, ibuprofen], histamine 2 receptor antagonists (H2RAs), cholesterol lowering medications), multivitamin use, and COVID-19 vaccination.

CI: confidence interval; COVID-19: coronavirus disease 2019; HR: hazard ratio; PPI: proton pump inhibitor.

**Supplementary Table S10.** Clinical characteristics of included participants after propensity score-matching

|  | **Regular PPI**  **user** | **Matched PPI**  **non-user** | **Overall** | **Standardized mean difference** |
| --- | --- | --- | --- | --- |
| **Number of participants, n (%)** | 9 940 (20%) | 39 760 (80%) | 49 700 (100%) |  |
| **Age, years, mean (SD)** | 59.4 (7.4) | 60.2 (7.0) | 60.1 (7.1) | **-0.11** |
| **Sex, female, n (%)** | 5 498 (55.3) | 22 400 (56.3) | 27 898 (56.1) | 0.02 |
| **Ethnicity, white, n (%)** | 9 518 (95.8) | 37 973 (95.5) | 47 491 (95.6) | 0.01 |
| **Deprivation index, mean (SD)** | -0.92 (3.3) | -0.76 (3.23) | -0.79 (3.24) | -0.05 |
| **Alcohol consumption, n (%)** |  |  |  | 0.06 |
| **Daily or almost daily** | 1 793 (18.0) | 6 985 (17.6) | 8 778 (17.7) |  |
| **3 or 4 times a week** | 1 913 (19.2) | 7 123 (17.9) | 9 036 (18.2) |  |
| **1 or 2 times a week** | 2 384 (24.0) | 9 182 (23.1) | 11 566 (23.3) |  |
| **1 to 3 times a month** | 1 173 (11.8) | 4 750 (11.9) | 5 923 (11.9) |  |
| **Special occasions only** | 1 510 (15.2) | 6 524 (16.4) | 8 034 (16.2) |  |
| **Never** | 1 156 (11.6) | 5 138 (12.9) | 6 294 (12.7) |  |
| **Smoking, n (%)** |  |  |  | 0.04 |
| **Never smoker** | 4 545 (45.7) | 17 368 (43.7) | 21 913 (44.1) |  |
| **Previous smoker** | 4 263 (42.9) | 17 754 (44.7) | 22 017 (44.3) |  |
| **Current smoker** | 1 132 (11.4) | 4 638 (11.7) | 5 770 (11.6) |  |
| **Physical activity, MET minutes/week, median (IQR)** | 1 527 (2 721.0) | 1 533 (2 494.5) | 1 530 (2 548.0) | 0.03 |
| **Fresh fruit intake, pieces, mean (SD)** | 2.0 (2.6) | 2.0 (2.6) | 2.0 (2.6) | 0.02 |
| **BMI, kg/m2, mean (SD)** | 29.2 (5.1) | 29.7 (5.6) | 29.6 (5.5) | 0.09 |
| **Indication of PPIs, n (%)** |  |  |  |  |
| **GERD** | 3 215 (32.3) | 3 996 (10.1) | 7211 (14.5) | **0.48** |
| **Peptic ulcer** | 554 (5.6) | 1 282 (3.2) | 1836 (3.7) | **0.10** |
| **Upper gastrointestinal bleeding** | 18 (0.2) | 38 (0.1) | 56 (0.1) | 0.02 |
| **Comorbidities, n (%)** |  |  |  |  |
| **Hypertension** | 4 083 (41.1) | 18 933 (47.6) | 23016 (46.3) | 0.13 |
| **Type 2 diabetes** | 124 (1.2) | 598 (1.5) | 722 (1.5) | 0.02 |
| **Renal failure** | 60 (0.6) | 216 (0.5) | 276 (0.6) | 0.01 |
| **Myocardial infarction** | 326 (3.3) | 1 472 (3.7) | 1798 (3.6) | 0.02 |
| **Stoke** | 135 (1.4) | 659 (1.7) | 794 (1.6) | 0.02 |
| **COPD** | 45 (0.5) | 175 (0.4) | 220 (0.4) | 0.001 |
| **Asthma** | 834 (8.4) | 3 489 (8.8) | 4 323 (8.7) | 0.01 |
| **Medication use, n (%)** |  |  |  |  |
| **Aspirin** | 2 429 (24.4) | 11 898 (29.9) | 14 327 (28.8) | **0.13** |
| **Non-aspirin NSAIDS** | 1 205(12.1) | 4 734 (11.9) | 5 939 (11.9) | 0.01 |
| **H2RA** | 295 (3.0) | 1 182 (3.0) | 1 477 (3.0) | 0.0003 |
| **Cholesterol lowering medication** | 1 526(15.4) | 7 311 (18.4) | 8 837 (17.8) | 0.08 |
| **Multivitamin use, n (%)** | 2 213(22.3) | 8 979 (22.6) | 11 192 (22.5) | 0.01 |

BMI: body mass index; COPD: chronic obstructive pulmonary disease; GERD: gastroesophageal reflux disease; H2RA: histamine 2 receptor antagonist; IQR: interquartile range; MET: metabolic equivalent of task; PPI: proton pump inhibitor; NSAIDS: non-steroidal anti-inflammatory drugs; SD: standard deviation.

**Supplementary Table S11.** Propensity score-matched analysis of associations of PPI use with the risk of influenza, pneumonia, COVID-19, and other respiratory infections

|  | **Cases / Person-years** | **Unadjusted**  **HR (95%CI)** | ***P*** | **Multivariable-adjusted model*** | ***P*** |
| --- | --- | --- | --- | --- | --- |
| **Influenza** |  |  |  |  |  |
| **Non-regular PPI use** | 560/1 582 | 1.00 (Reference) |  | 1.00 (Reference) |  |
| **Regular PPI use** | 183/539 | 1.31 (1.11-1.55) | **0.001** | 1.33 (1.12-1.58) | **0.001** |
| **Pneumonia** |  |  |  |  |  |
| **Non-regular PPI use** | 1 263/5 688 | 1.00 (Reference) |  | 1.00 (Reference) |  |
| **Regular PPI use** | 377/1 699 | 1.20 (1.07-1.35) | **0.001** | 1.33 (1.18-1.50) | **<0.001** |
| **COVID-19** |  |  |  |  |  |
| **Non-regular PPI use** | 5 442/6 463 | 1.00 (Reference) |  | 1.00 (Reference) |  |
| **Regular PPI use** | 1 429/1 689 | 1.05 (0.97-1.14) | 0.226 | 1.07 (0.98-1.17) | 0.128 |
| **Other upper respiratory infection** |  |  |  |  |  |
| **Non-regular PPI use** | 4 028/14 496 | 1.00 (Reference) |  | 1.00 (Reference) |  |
| **Regular PPI use** | 1 111/3 964 | 1.20 (1.12-1.28) | **<0.001** | 1.18 (1.10-1.26) | **<0.001** |
| **Other lower respiratory infection** |  |  |  |  |  |
| **Non-regular PPI use** | 4 849/18 589 | 1.00 (Reference) |  | 1.00 (Reference) |  |
| **Regular PPI use** | 1 475/5 549 | 1.30 (1.22-1.37) | **<0.001** | 1.33 (1.26-1.42) | **<0.001** |

*Adjusted for age, gastroesophageal reflux disease, peptic ulcer, and aspirin use.

CI: confidence interval; COVID-19: coronavirus disease 2019; HR: hazard ratio; PPI: proton pump inhibitor.

**Supplementary Table S12.** Propensity score-matched analysis of associations of PPI use with COVID-19 severity and mortality.

|  | **Cases / Person-years** | **Unadjusted**  **HR (95%CI)** | ***P*** | **Multivariable-adjusted model*** | ***P*** |
| --- | --- | --- | --- | --- | --- |
| **COVID-19 severity** |  |  |  |  |  |
| **Non-regular PPI use** | 490/373 | 1.00 (Reference) |  | 1.00 (Reference) |  |
| **Regular PPI use** | 138/100 | 1.10 (0.91-1.34) | 0.303 | 1.25 (1.02-1.52) | **0.028** |
| **COVID-19 mortality** |  |  |  |  |  |
| **Non-regular PPI use** | 163/112 | 1.00 (Reference) |  | 1.00 (Reference) |  |
| **Regular PPI use** | 48/36 | 1.15 (0.84-1.59) | 0.389 | 1.45 (1.04-2.02) | **0.030** |

*Adjusted for age, gastroesophageal reflux disease, peptic ulcer, and aspirin use.

CI: confidence interval; COVID-19: coronavirus disease 2019; HR: hazard ratio; PPI: proton pump inhibitor.

**Supplementary Table S13.** Comparisons between proton pump inhibitor (PPI) and histamine-2 receptor antagonist (H2RA) users for COVID-19 severity and mortality.

|  | **Cases / Person-years** | **HR (95% Cl)*** | ***P*** |
| --- | --- | --- | --- |
| **COVID-19 severity** |  |  |  |
| **Regular H2RA use** | 42/35 | 1.00 (Reference) |  |
| **Regular PPI use** | 136/99 | 0.91 (0.64-1.30) | 0.608 |
| **COVID-19 mortality** |  |  |  |
| **Regular H2RA use** | 14/10 | 1.00 (Reference) |  |
| **Regular PPI use** | 48/36 | 0.83 (0.45-1.56) | 0.745 |

CI: confidence interval; COVID-19: coronavirus disease 2019; HR: hazard ratio; PPI: proton pump inhibitor.

*Adjusted for age, sex, ethnicity, deprivation index, smoking, alcohol consumption, physical activity, fresh fruit intake, body mass index, any indication of PPIs (gastroesophageal reflux disease [GERD], peptic ulcer, upper gastrointestinal bleeding), comorbidities (hypertension, type 2 diabetes, renal failure, myocardial infarction, stroke, chronic obstructive pulmonary disease [COPD], asthma), medications (aspirin, non-aspirin non-steroidal anti-inflammatory drugs [NSAIDs, ibuprofen], cholesterol lowering medications), multivitamin use, and COVID-19 vaccination (for COVID-19-related outcomes).
